# Gargle sample is an effective option in a novel fully automated molecular point-of-care test for influenza: a multicenter study

**DOI:** 10.1101/2022.06.03.22275936

**Authors:** Norihito Kaku, Tomohito Urabe, Tetsuya Iida, Chyuns Yun, Yoshiyuki Nishida, Yasunori Onitsuka, Kohji Hashiguchi, Kiyoto Hirose, Akimitsu Tomonaga, Koichi Izumikawa, Hiroshi Mukae, Katsunori Yanagihara

## Abstract

**Background:** We conducted a multicenter study to evaluate the performance of a novel fully automated molecular point-of-care test using transcription-reverse transcription concerted reaction that can detect influenza A and B within 15 minutes in nasopharyngeal swabs and gargle samples (TRCsatFLU).

**Methods:** Patients who visited or were hospitalized at eight clinics and hospitals with influenza-like illnesses between December 2019 and March 2020 participated in this study. We collected nasopharyngeal swabs from all patients and gargle samples from patients whom the physician judged fit to perform gargling. The result of TRCsatFLU was compared to a conventional reverse transcription-polymerase chain reaction (RT-PCR). If the results of TRCsatFLU and conventional RT-PCR were different, the samples were analyzed by sequencing.

**Results:** We evaluated 233 nasopharyngeal swabs and 213 gargle samples from 244 patients.. The average age of the patients was 39.3 ± 21.2. Of the patients, 68.9% visited a hospital within 24 h of symptom onset. The most common symptoms were fever (93.0%), fatigue (79.5%), and nasal discharge (64.8%). All patients in whom the gargle sample was not collected were children. Influenza A or B was detected in 98 and 99 patients in nasopharyngeal swabs and gargle samples using TRCsatFLU, respectively. Four and five patients in nasopharyngeal swabs and gargle samples, respectively, with different TRCsatFLU and conventional RT-PCR results. Influenza A or B was detected using sequencing in all samples with different results. Based on the combined conventional RT-PCR and sequencing results, the sensitivity, specificity, positive predictive value (PPV), and negative predictive value (NPV) of TRCsatFLU for influenza detection in nasopharyngeal swabs were 0.990, 1.000, 1.000, and 0.993, respectively. In the gargle samples, the sensitivity, specificity, PPV, and NPV of the TRCsatFLU for detecting influenza were 0.971, 1.000, 1.000, and 0.974, respectively.

**Conclusions:** The TRCsatFLU showed great sensitivity and specificity for the detection of influenza in nasopharyngeal swabs and gargle samples.

**Trial registration:** This study was registered in the UMIN Clinical Trials Registry (reference number: UMIN000038276) on October 11, 2019. Before sample collection, written informed consent for the participation and publication of this study was obtained from all participants.

## Background

Influenza is a major infectious disease that spreads during the winter with clinics receiving many influenza patients during the flu season. As antiviral agents for influenza, such as neuraminidase inhibitors are already available, it is essential to accurately test and diagnose influenza. Rapid influenza diagnostic tests (RIDTs) have been widely used in clinics because of their simplicity and speed. However, they often have low sensitivity, especially in the early stage of influenza, compared to nucleic acid tests. [1] Therefore, molecular point-of-care (POC) tests and highly sensitive automated immunochromatographic antigen tests (digital immunoassays, DIAs) for influenza have been developed and showed markedly higher sensitivities than traditional rapid influenza diagnostic tests.[2] Recently, applying transcription-reverse transcription concerted reaction (TRC), a novel fully automated molecular POC machine (TRCsat®; Tosoh, Tokyo, Japan) with a dedicated single-use cartridge for influenza (TRCsatFLU; Tosoh, Tokyo, Japan) was developed. TRC is a gene-detecting method that involves rapid isothermal RNA amplification using an intercalation-activating fluorescence (INAF) probe, and It has been used to diagnose tuberculosis, nontuberculous mycobacterial infections, mycoplasma pneumonia, chlamydial infections, and gonorrhea. [3,4] TRCsatFLU contains all the elements required for detecting influenza A and B, and it can detect influenza A and B within 15 min. One feature of TRCsatFLU, apart from other POC molecular tests, is that it can detect influenza from gargle samples in addition to nasopharyngeal swabs with simple sample preparation without purification. In a previous single-center study, TRCsatFLU showed comparable performance to the conventional reverse transcription-polymerase chain reaction (RT-PCR) method for detecting influenza viruses in nasopharyngeal swabs and gargle samples obtained from patients with influenza-like illness (ILI). [5] However, in a previous study, we divided the study period into two and collected gargle samples only from the second period. In addition, because the study was conducted in only a secondary hospital where many adult patients with underlying diseases were seen, it is unclear whether TRCsatFLU could show the same results in patients with ILI, including children, who visit clinics. Therefore, we conducted a multicenter study in several clinical settings, including pediatric clinics, to evaluate the performance of TRCsatFLU for detecting influenza viruses in nasopharyngeal swabs and gargle samples.

## Methods

### Study design

This prospective observational study was conducted between December 16, 2019 and March 25, 2020. The original plan was to collect samples until May 31, 2022 but the study was suspended early on March 25, 2020 when the spread of coronavirus disease 2019 (COVID-19) began in Japan. [6–8] Due to the continued COVID-19 pandemic, the study was not resumed, and the analysis was conducted on the samples collected by March 25, 2020. We selected seven internal medicine clinics, pediatrics, and otorhinolaryngology clinics in Nagasaki Prefecture that could participate, the Urabe Otorhinolaryngology Clinic, Iida Naika Syounika Clinic, Ohisama Pediatric Clinic, Nishida Gastrointestinal Intermedicine Clinic, Onitsuka Internal Medicine Clinic, Hirose Clinic, and Tomonaga Medical Clinic. We also included a hospital that participated in the previous study, the Japanese Red Cross Nagasaki Genbaku Hospital. In eight medical facilities, we included patients who visited or were hospitalized with influenza-like illness (ILI), as defined by the World Health Organization’s case definition.[9] Patients were excluded if they were administered anti-influenza agents within one month before sampling. After obtaining informed consent, nasopharyngeal swabs and gargle samples were collected. Two nasopharyngeal swabs (1PY1502P; Japan Cotton Swab Industry, Limited, Tokyo, Japan) were collected from all the patients by a healthcare provider. Gargle samples were collected from patients whom the physician judged to be able to perform gargling. In gargle samples, the patients gargled for 5 seconds with 20 mL of water (water for injection; Hikari Pharmaceutical CO., LTD. Tokyo, Japan), which was collected. Gargle samples were stored at -20°C in a container (Multi-purpose container, 70 mL; Sarstedt, K.K., Tokyo, Japan) until further analysis. One of the swabs was used in each medical facility for detecting influenza by DIAs using silver amplification immunochromatography (FUJI DRI-CHEM IMMUNO AG Cartridge FluAB; Fujifilm, Kanagawa, Japan)[10], according to manufacturer’s instruction. Another nasopharyngeal swab and gargle samples were stored at -20°C in a sealable tube (PP screw cap test tube; Maruemu Corporation, Osaka, Japan) without media until further analysis. The physicians determined the clinical diagnosis based on medical history, physical findings, and DIAs results, from which they produced a clinical report for each patient. Since TRCsatFLU was not approved in Japan when this study was conducted, and it was necessary to prevent the use of TRCsatFLU results for the diagnosis of influenza at medical facilities, nasopharyngeal swabs and gargle samples were transferred to Tosoh Corporation for performing TRCsatFLU and RT-PCR. All information, such as clinical report forms and TRCsatFLU and RT-PCR results, was summarized and analyzed at Nagasaki University Hospital. If the results of TRCsatFLU were different from those of RT-PCR, the samples were analyzed by sequencing at Tosoh Corporation.

### Ethics

This study was approved by the ethics committee of Nagasaki University Hospital (approval number:19121603) and registered in the UMIN Clinical Trials Registry (reference number: UMIN000038276). Before sample collection, written informed consent for the participation and publication of this study was obtained from all participants.

### Data collection

To compile data on patient characteristics, we collected information on sex, age, underlying diseases, history of influenza vaccination, time since onset of symptoms, body temperature at the time of consultation, clinical diagnosis, results of DIAs, treatment for ILI, and the following signs and symptoms: fever (body temperature ≥37.5 °C), cough, sore throat, nasal discharge, headache, arthralgia and myalgia, fatigue, nausea, and diarrhea.

### TRC

For TRCsatFLU, we soaked a new swab in a gargle sample for 5 sec, and the swab containing the gargle solution was mixed with 1 mL extraction buffer containing surfactant, and it was infected into a single-use cartridge that contains all the elements required for rapid TRC. Nasopharyngeal swabs were mixed with 1 mL extraction buffer containing surfactant. Of the extraction buffer, 140 μL was aliquoted for RT-PCR, and the remaining 860 μL was injected into the TRCsatFLU cartridge. Next, the cartridge was set in TRCsat®, and nucleic acid amplification, detection, and determination of results were automatically performed in the instrument. The procedure of rapid TRC in TRCsat® is as follows: the samples were incubated at 52 °C for 1 min and then mixed 30 μL of the sample with a dry reagent containing enzymes, substrates, primers, and INAF probes and incubated at 46 °C to monitor fluorescence; it was automatically determined as positive when the fluorescence intensity ratio of the reaction solution exceeded 1.2. TRCsatFLU was designed to detect Influenza A(H1N1), A(H1N1)pdm09, A (H3N2), and B (Victoria and Yamagata lineages), but TRCsat® only showed the results with influenza A or B positive or influenza negative.

### RT-PCR and sequencing

We performed RNA extraction and RT-PCR according to the Influenza Diagnosis Manual 4th edition (National Institute of Infectious Diseases, 2019).[11] In gargle samples, total RNA was isolated from 140 μL of a gargle sample using a QIAamp Viral RNA Mini Kit (Qiagen, Hilden, Germany). For nasopharyngeal swabs, total RNA was isolated from 140 μL of the extraction buffer using a QIAamp Viral RNA Mini Kit (Qiagen, Hilden, Germany). RT-PCR was performed using the primers and probes listed in Table S1, the One-Step PrimeScript™ RT-PCR Kit (TAKARA BIO, Shiga, Japan), and QuantStudio5 Real-Time PCR System (Thermo Fisher Scientific, Massachusetts, USA). In brief, 2 μL of extracted RNA was added to 10 μL of 2X One Step RT-PCR Buffer III, 0.4 μL each of 10 μM primers (Table S1), 0.25μL of 10.2 μM Taqman Probe, 0.4 μL of 50X ROX Reference Dye II, 0.4 μL of PrimeScript RT enzyme Mix II, 0.4 μL of TaKaRa Ex Taq HS, and 5.75 μL of RNase free water. The conditions consisted of 1 cycle of 5 min at 42 °C, 10 sec at 95 °C and followed by 45 cycles of 5 s at 95 °C, 34 s at 55 °C for H1N1 or 58 °C for H3N2 and B. The result was analyzed using QuantStudio (Thermo Fisher Scientific, Massachusetts, USA), in which a cycle threshold value (Ct-value)□<□40 was defined as a positive result.

If the results of TRCsatFLU were different from those of RT-PCR, the TRC and RT-PCR products were analyzed by sequencing according to the Influenza Diagnosis Manual 4th edition (National Institute of Infectious Diseases, 2019).[11] In brief, positive samples were purified using QIAquick PCR Purification kit (Qiagen, Hilden, Germany). Sequencing employed the ABI Big Dye Terminator system (ThermoFisher Scientific, Waltham, MA, USA). It was performed at a contract sequencing facility (FASMAC Co., Ltd. Kanagawa, Japan). For each sequencing reaction, 50 ng template and 3.2 pmol primers (Table S1) were used.

### Statistical analysis

All statistical analyses were performed using R (the R Foundation for Statistical Computing, Vienna, Austria; version 4.0.3). Fisher’s exact test was used to compare categorical variables and the Mann–Whitney U test was used to compare continuous variables. The statistical significance level was set at p <0.05. The sensitivity (Se), specificity (Sp), positive predictive value (PPV), and negative predictive value (NPV) of the TRCsatFLU against the combined results of RT-PCR and sequencing were calculated with 95% confidence intervals (95% CI) as in the previous study. [5]

## Results

### Patient characteristics

During the study period, 286 patients with ILI participated. Excluded patients and samples were shown in Fig1: one patient was excluded due to withdrawal of consent; 11 nasopharyngeal swabs were excluded due to testing protocol deviation that mixed nasopharyngeal swabs with 1mL extraction buffer for 10 sec, not 5 sec; 40 patients were excluded due to failure of the freezer at the clinic where the samples were stored. Finally, 233 nasopharyngeal swabs and 213 gargle samples obtained from 244 patients were evaluated. Patient characteristics are shown in Table 1. The average age of the patients was 39.3 ± 21.2. The proportion of patients with underlying diseases and a history of influenza vaccination was 33.2% and 50.0%, respectively. Of the patients, 68.9% visited a hospital within 24 h of symptom onset. The most common symptoms were fever (93.0%), fatigue (79.5%), and nasal discharge (64.8%). In the clinical diagnosis based on history, physical findings, and the results of DIAs, 44.3% of the patients were diagnosed with influenza and 43.0% were diagnosed with acute upper respiratory infection. All patients in whom gargle samples were not collected were children. The average age of the patients was 2.9, and most of them (26/31) were under the age of 5.

**Figure 1.**
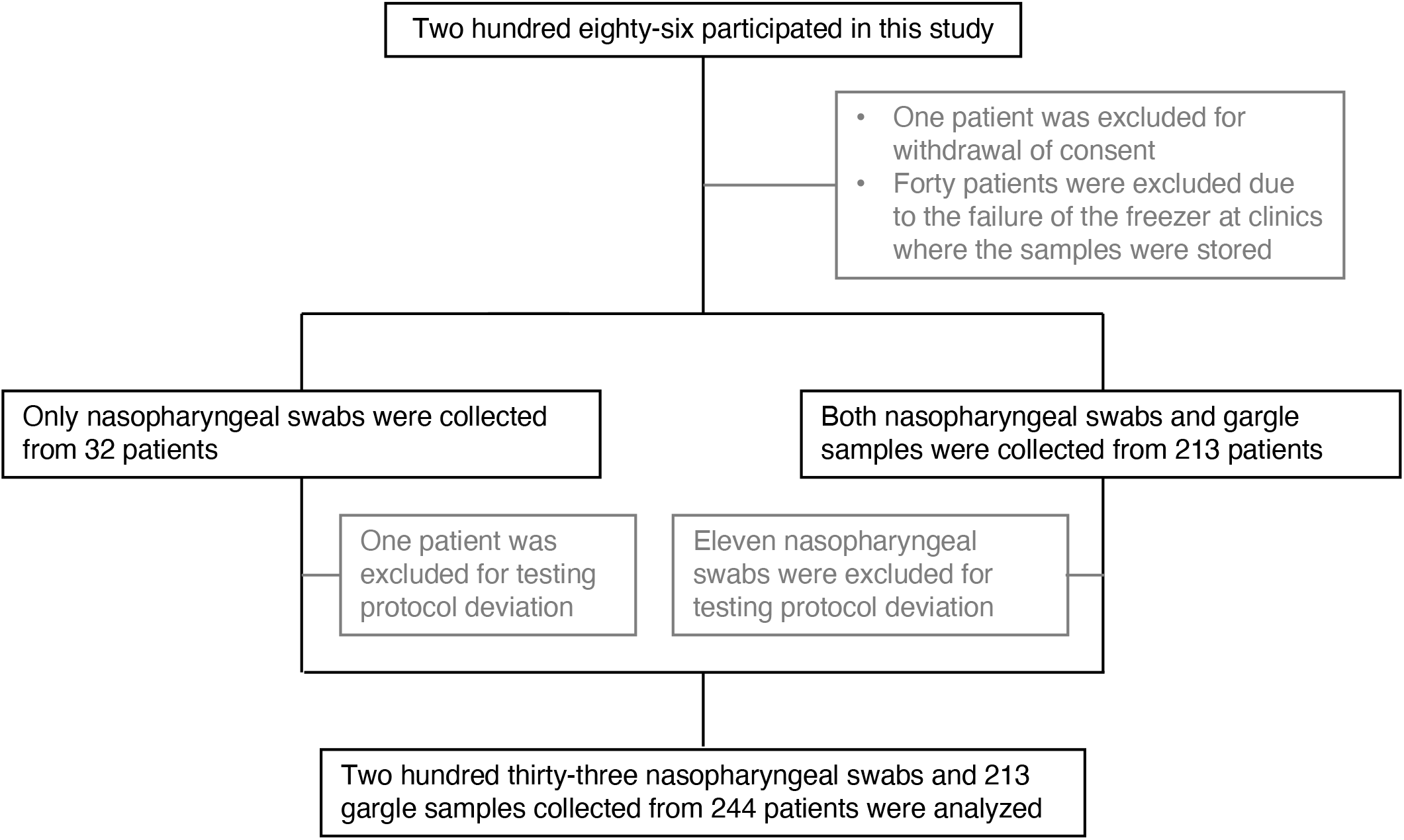
Participant flow diagram. Participant flow diagram showing progression through phases of prospective observational study.

**Table 1.** Patient characteristics.

| Characteristic | All | Nasopharyngeal<br>swabs and gargle<br>samples | Nasopharyngeal<br>swabs only | Gargle samples<br>only |
| --- | --- | --- | --- | --- |
|  | N=244 | N=202 | N=31 | N=11 |
|  | N (%) | N (%) | N (%) | N (%) |
| Age (average ± S.D.) | 39.3 ± 21.2 | 44.6 ± 16.6 | 2.9 ± 2.9 | 43.5 ± 24.0 |
| Under 16 years old | 37 (15.2%) | 4 (2.0%) | 31 (100%) | 2 (18.2%) |
| Gender, female | 138 (56.6%) | 119 (58.9%) | 14 (45.2%) | 5 (45.5%) |
| Underlying diseases | 81 (33.2%) | 75 (37.1%) | 3 (9.7%) | 3 (27.3%) |
| History of |  |  |  |  |
| Influenza vaccination | 122 (50.0%) | 102 (50.5%) | 14 (45.2%) | 6 (54.5%) |
| Time since onset of symptoms |  |  |  |  |
| 0 - 6 h | 33 (13.5%) | 26 (12.9%) | 5 (16.1%) | 2 (18.2%) |
| 6 - 12 h | 31 (12.7%) | 21 (10.4%) | 9 (29.0%) | 1 (9.1%) |
| 12 - 24 h | 104 (42.6%) | 86 (42.6%) | 13 (41.9%) | 5 (45.5%) |
| 24 - 48 h | 45 (18.4%) | 41 (20.3%) | 3 (9.7%) | 1 (9.1%) |
| 48 - 72 h | 21 (8.6%) | 20 (9.9%) | 1 (9.7%) | 0 (0.0%) |
| 72 h - | 7 (2.9%) | 6 (3.0%) | 0 (0.0%) | 1 (9.1%) |
| Unknown | 3 (1.2%) | 2 (1.0%) | 0 (0.0%) | 1 (9.1%) |
| Symptoms |  |  |  |  |
| Fever | 227 (93.0%) | 186 (92.1%) | 31 (100%) | 10 (90.9%) |
| Fatigue | 194 (79.5%) | 177 (87.6%) | 1 (3.2%) | 8 (72.7%) |
| Nasal discharge | 158 (64.8%) | 128 (63.4%) | 20 (64.5%) | 10 (90.9%) |
| Cough | 157 (64.3%) | 137 (67.8%) | 18 (58.1%) | 2 (18.2%) |
| Headache | 152 (62.3%) | 144 (71.3%) | 1 (3.2%) | 7 (63.6%) |
| Sore throat | 150 (61.5%) | 141 (69.8%) | 2 (6.5%) | 7 (63.6%) |
| Arthralgia | 129 (52.9%) | 124 (61.4%) | 0 (0.0%) | 5 (45.5%) |
| Myalgia | 97 (39.8%) | 92 (45.5%) | 9 (29.0%) | 4 (36.4%) |
| Diarrhea | 15 (6.1%) | 14 (6.9%) | 1 (3.2%) | 1 (9.1%) |
| Nausea | 13 (5.3%) | 11 (5.4%) | 0 (0.0%) | 1 (9.1%) |
| Results of influenza antigen test using silver amplification immunochromatography |  |  |  |  |
| Influenza A | 93 (38.1%) | 84 (41.6%) | 5 (16.1%) | 4 (36.4%) |
| Influenza B | 1 (0.4%) | 1 (0.5%) | 0 (0.0%) | 0 (0.0%) |
| Negative | 150 (61.5%) | 117 (57.9%) | 26 (83.9%) | 7 (63.6%) |
| Clinical Diagnosis |  |  |  |  |
| Influenza | 108 (44.3%) | 98 (48.5%) | 6 (19.4%) | 4 (36.4%) |
| Acute upper respiratory infection | 105 (43.0%) | 77 (38.1%) | 21 (67.7%) | 7 (63.6%) |
| Acute bronchitis | 17 (7.0%) | 14 (6.9%) | 3 (9.7%) | 0 (0.0%) |
| Others | 14 (5.7%) | 13 (6.4%) | 1 (3.2%) | 0 (0.0%) |

S.D., standard deviation

### Detection of influenza by TRCsatFLU in nasopharyngeal swabs and gargle samples

The results of the TRCsatFLU and RT-PCR are shown in Table 2. Influenza was detected in nasopharyngeal swabs and gargle samples using TRCsatFLU in 41.6% and 46.5% of patients, respectively. The positive rate for influenza was highest at 24–48 h after the onset of symptoms in both types of specimens (Fig. 2). Compared to DIAs, the positive rate for influenza within 6 h after the onset of symptoms in TRCsatFLU and RT-PCR in nasopharyngeal swabs was two times higher than that in DIAs (Fig. 2A). There were 14 cases with a clinical diagnosis of influenza despite negative DIAs; influenza was detected in 5 of 14 nasopharyngeal swabs and 4 of 13 gargle samples by TRCsatFLU (Table S2). Additionally, influenza was detected by TRCsatFLU in 6 nasopharyngeal swabs and 10 gargle samples collected from the patients diagnosed as other than influenza (Table S2).

**Table 2.** Results of TRCsatFLU.

|  | Nasopharyngeal swabs | Gargle samples |
| --- | --- | --- |
|  | (N=233) | (N=213) |
|  | N (%) | N (%) |
| TRC<sub>sat</sub>FLU |  |  |
| Influenza A | 97 (41.6%) | 99 (46.5%) |
| Influenza B | 1 (0.4%) | 0 (0.0%) |
| Negative | 135 (57.9%) | 114 (53.5%) |
| RT-PCR |  |  |
| Influenza A(H1N1)pdm09 | 96 (41.2%) | 100 (46.9%) |
| Influenza B | 0 (0.0%) | 0 (0.0%) |
| Negative | 137 (58.8%) | 113 (53.1%) |

**Figure 2.**
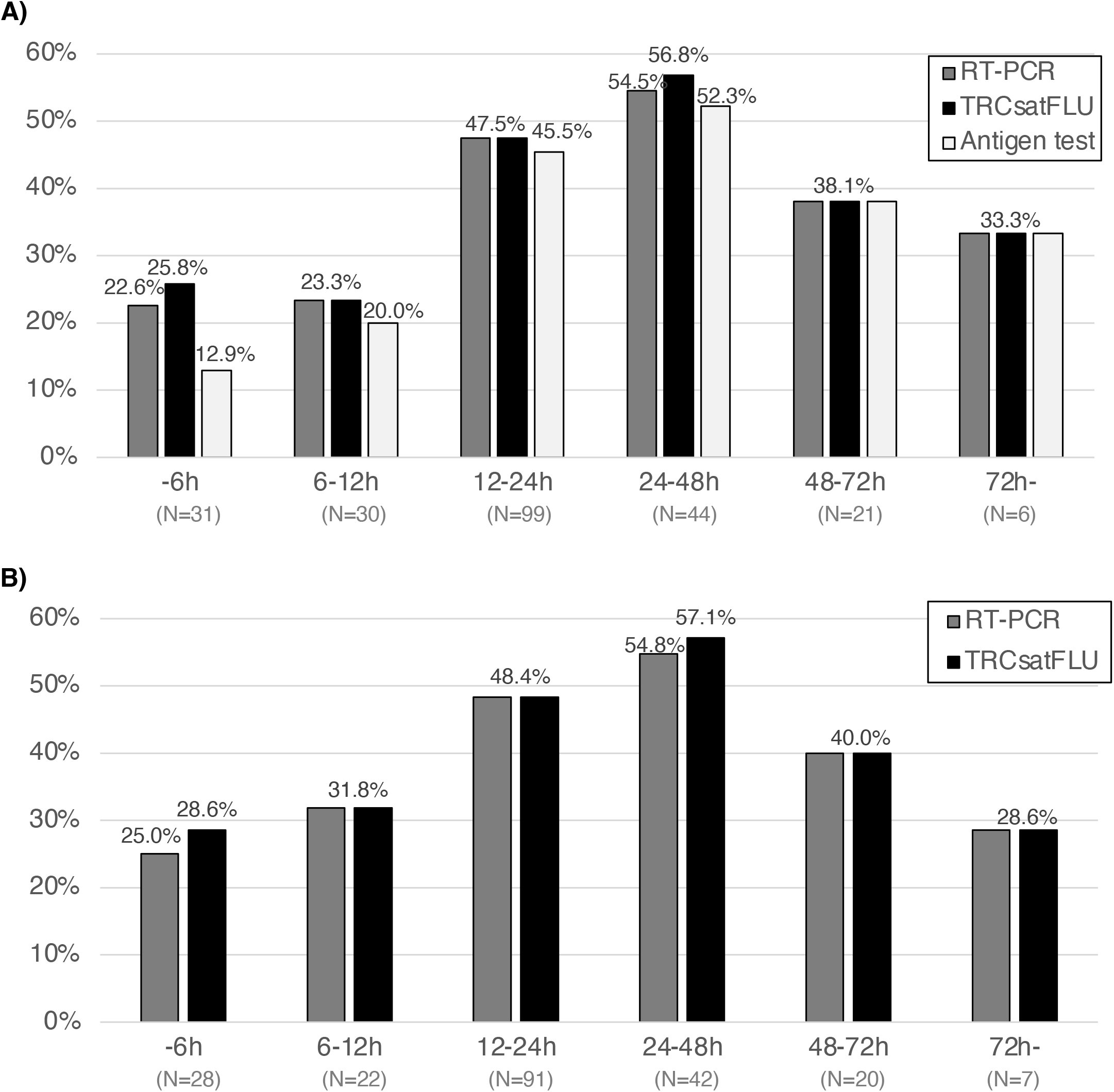
Positive rate of influenza in each test by time since onset of symptoms. Nasopharyngeal swabs were collected from all patients using two swabs (A). Gargle samples were collected from patients whom the physician judged fit to perform gargling (B). A highly sensitive automated antigen test was performed using silver amplification immunochromatography for influenza (FUJI DRI-CHEM IMMUNO AG Cartridge FluAB; Fujifilm, Kanagawa, Japan) at clinics and hospitals. Nasopharyngeal swabs and gargle samples for TRCsatFLU and RT-PCR were stored at −20 °C until further analysis. The samples were transferred to Tosoh Corporation to perform TRCsatFLU and RT-PCR.

### Performance of TRCsatFLU on the detection of influenza

Four nasopharyngeal swabs showed different results in RT-PCR and TRCsatFLU: one tested positive for influenza A(H1N1)pdm09 only with RT-PCR; two and one tested positive for influenza A and B, respectively, only with TRCsatFLU. In those four samples, influenza B and influenza A(H1N1)pdm09 were detected by sequencing in one and three samples, respectively. Based on the combined RT-PCR and sequencing results, the Se, Sp, PPV, and NPV of TRCsatFLU in nasopharyngeal swabs were 0.990, 1.000, 1.000, and 0.993, respectively (Table 3).

**Table 3.** Performance of TRCsatFLU in nasopharyngeal swabs and gargle samples.

|  | Nasopharyngeal swabs | Gargle samples |
| --- | --- | --- |
|  | (N=233) | (N=213) |
| True-positive | 98 | 99 |
| True-negative | 134 | 111 |
| False-positive | 0 | 0 |
| False-negative | 1 | 3 |
| Se (95% CI) | 0.990 (0.945-1.000) | 0.971 (0.916-0.994) |
| Sp (95% CI) | 1.000 (0.959-1.000) | 1.000 (0.951-1.000) |
| PPV (95% CI) | 1.000 (0.945-1.000) | 1.000 (0.951-1.000) |
| NPV (95% CI) | 0.993 (0.959-1.000) | 1.000 (0.925-0.995) |

CI, confidence

Five gargle samples showed different results in RT-PCR and TRCsatFLU: three tested positives for influenza A(H1N1)pdm09 only with RT-PCR, and two tested positive for influenza A only with TRCsatFLU. Influenza A(H3N2) was detected by sequencing in one of the samples that tested positive only with TRCsatFLU, and influenza A(H1N1)pdm09 was detected in all other samples. Based on the combined RT-PCR and sequencing results, the Se, Sp, PPV, and NPV of TRCsatFLU in nasopharyngeal swabs were 0.971, 1.000, 1.000, and 0.974, respectively (Table 3).

### Comparison of TRCsatFLU results between nasopharyngeal swabs and gargle samples

We evaluated TRCsatFLU results in nasopharyngeal swabs and gargle samples collected from the same patients. Of 202 patients, 86 were positive for influenza in both specimens. However, 14 had different results between nasopharyngeal swabs and gargle samples: seven tested positive for influenza A only in nasopharyngeal swabs, one tested positive for influenza B only in nasopharyngeal swabs, and six patients tested positive for influenza A only in gargle samples.

In the patients who tested positive for influenza only in nasopharyngeal swabs, the percentage of fever and headache, the percentage of diagnosing influenza, the influenza-positive rate by DIAs and RT-PCR in nasopharyngeal swabs, and the influenza-positive rate by RT-PCR in gargle samples were significantly lower than the patients who tested positive for influenza in both samples (Table 4). In the patients who tested positive for influenza only in gargle samples, the percentage of patients who visited the medical facilities within 6 h since the onset of symptoms was significantly higher than those who tested positive for influenza in both samples (Table 4). Additionally, the percentage of diagnosing influenza, the influenza-positive rate by DIAs and RT-PCR in nasopharyngeal swabs were significantly lower in the patients who tested positive for influenza only in gargle samples than those in the patients who tested positive for influenza in both samples (Table 4). However, there was no difference between the patients who tested positive for influenza only in nasopharyngeal swabs and only in gargle samples.

**Table 4.** Characteristics of patients with different results between nasopharyngeal swabs and gargle sample

|  | Positive in both samples<br>(N=86) |  | Positive only in nasopharyngeal swabs<br>(N=8) |  | *P* value compared to positive in both samples | Positive only in gargle samples<br>(N=6) |  | *P* value compared to positive in both samples | *P* value compared positive only in nasopharyngeal swabs and only in gargle samples |
| --- | --- | --- | --- | --- | --- | --- | --- | --- | --- |
|  | N | (%) | N | (%) |  | N | (%) |  |  |
| Age (average ± S.D) | 44.9 | ± 22.9 | 50.9 | ± 22.9 | NS | 41.3 | ± 10.9 | NS | NS |
| Under 16 years old | 3 | (3.5%) | 1 | (12.5%) | NS | 0 | (0.0%) | NS | NS |
| Gender, female | 53 | (61.6%) | 5 | (62.5%) | NS | 1 | (16.7%) | 0.078 | 0.138 |
| Underlying diseases | 29 | (33.7%) | 4 | (50.0%) | NS | 2 | (33.3%) | NS | NS |
| History of Influenza vaccination | 33 | (38.4%) | 4 | (50.0%) | NS | 2 | (33.3%) | NS | NS |
| Time since onset of symptoms |  |  |  |  |  |  |  |  |  |
| 0–6 h | 6 | (7.0%) | 2 | (25.0%) | 0.137 | 3 | (50.0%) | 0.011 | NS |
| 6–12 h | 6 | (7.0%) | 1 | (12.5%) | NS | 0 | (0.0%) | NS | NS |
| 12–24 h | 43 | (50.0%) | 1 | (12.5%) | 0.063 | 3 | (50.0%) | NS | 1 |
| 24–48 h | 21 | (24.4%) | 3 | (37.5%) | NS | 0 | (0.0%) | NS | 0.245 |
| 48–72 h | 7 | (8.1%) | 1 | (12.5%) | NS | 0 | (0.0%) | NS | 0.209 |
| 72+ h | 2 | (2.3%) | 0 | (0.0%) | NS | 0 | (0.0%) | NS | NS |
| Unknown | 1 | (1.2%) | 0 | (0.0%) | NS | 0 | (0.0%) | NS | NS |
| Symptoms |  |  |  |  |  |  |  |  |  |
| Fever | 82 | (95.3%) | 5 | (62.5%) | 0.012 | 6 | (100%) | NS | 0.209 |
| Fatigue | 77 | (89.5%) | 8 | (100%) | NS | 5 | (83.3%) | NS | NS |
| Nasal discharge | 54 | (62.8%) | 5 | (62.5%) | NS | 3 | (50.0%) | NS | NS |
| Cough | 70 | (81.4%) | 8 | (100%) | NS | 5 | (83.3%) | NS | NS |
| Headache | 70 | (81.4%) | 4 | (50.0%) | 0.060 | 5 | (83.3%) | NS | NS |
| Sore throat | 59 | (68.6%) | 7 | (87.5%) | NS | 5 | (83.3%) | NS | NS |
| Arthralgia | 58 | (67.4%) | 5 | (62.5%) | NS | 4 | (66.7%) | NS | NS |
| Myalgia | 48 | (55.8%) | 4 | (50.0%) | NS | 1 | (16.7%) | 0.093 | NS |
| Diarrhea | 5 | (5.8%) | 0 | (0.0%) | NS | 1 | (16.7%) | NS | NS |
| Nausea | 4 | (4.7%) | 0 | (0.0%) | NS | 0 | (0.0%) | NS | NS |
| Results of DIAs |  |  |  |  |  |  |  |  |  |
| Influenza A | 80 | (93.0%) | 3 | (37.5%) | <0.001 | 1 | (16.7%) | <0.001 | NS |
| Negative | 6 | (7.0%) | 5 | (62.5%) |  | 5 | (83.3%) |  |  |
| Clinical Diagnosis |  |  |  |  |  |  |  |  |  |
| Influenza | 83 | (96.5%) | 5 | (62.5%) | 0.007 | 2 | (33.3%) | <0.001 | 0.138 |
| Acute upper respiratory infection | 3 | (3.5%) | 1 | (12.5%) | NS | 3 | (50.0%) | 0.003 | 0.245 |
| Acute bronchitis | 0 | (0.0%) | 2 | (25.0%) | 0.006 | 1 | (16.7%) | 0.065 | NS |
| Results of RT-PCR in nasopharyngeal swabs |  |  |  |  |  |  |  |  |  |
| Influenza A(H1N1)pdm09 | 85 | (98.8%) | 6 | (75.0%) | 0.018 | 1 | (16.7%) | <0.001 | 0.103 |
| Negative | 1 | (1.1%) | 2 | (25.0%) |  | 5 | (83.3%) |  |  |
| Results of RT-PCR in gargle samples |  |  |  |  |  |  |  |  |  |
| Influenza A(H1N1)pdm09 | 85 | (98.8%) | 2 | (25.0%) | <0.001 | 5 | (83.3%) | 0.127 | 0.103 |
| Negative | 1 | (1.1%) | 6 | (75.0%) |  | 1 | (16.7%) |  |  |

NS, not significant (P > 0.2)

## Discussion

TRCsatFLU showed greater sensitivity and specificity in both nasopharyngeal swabs and gargle samples compared to the combined RT-PCR and sequencing. The sensitivity/specificity of the TRCsatFLU in nasopharyngeal swabs and gargle samples were 0.990/1.000 and 0.971/1.000, respectively, which are similar to those of the previous single-center study (1.000/1.000 and 0.946/1.000, respectively). [5] We conducted this study by adding seven clinics to the hospital that participated in the previous study. Therefore, there were differences in patient backgrounds between this study and the previous study. The average age in this study was ten years younger than that in the previous study because only this study included children. In addition, the percentages of patients with symptoms such as fever, fatigue, nasal discharge, headache, sore throat, arthralgia, and myalgia in this study were higher than those in the previous study. Fewer patients visited clinics and hospitals 72 hours after symptom onset compared to those in the previous study (2.9% versus 13.8%), which might have influenced the difference. The proportion of influenza variants was also different between the two studies. In the previous study, the most detected variant was influenza B, and the most detected influenza A variant was. In contrast, in this study, the most detected variant was influenza A(H1N1)pdm09, and influenza B and A(H3N2)weres detected from only one patient each. Thus, despite differences in patient backgrounds and influenza variants, the TRCsatFLU showed the same test performance in both studies.

TRCsatFLU can detect influenza A and B within 15 min. The most common RIDTs are antigen tests using immunochromatography in nasopharyngeal swabs. However, since conventional antigen tests have very low sensitivity in the early stages of influenza, [1] in this study, commercially available high-sensitivity DIAs using silver amplification immunochromatography [10] were used at clinics and hospitals for clinical diagnosis. However, the positive rate for influenza in DIAs was obviously lower than that of TRCsatFLU in this study. Notably, in patients who visited clinics or hospitals within 6 hours from the onset of symptoms, the positive rate for influenza in DIAs was half of that for TRCsatFLU. In addition, there were 14 cases with a clinical diagnosis of influenza despite negative DIAs. Among them, influenza was detected in 5 of 14 nasopharyngeal swabs and 4 of 13 gargle samples by TRCsatFLU. Those results are similar to the results of a multicenter study that we conducted at the same time as this study. In the study, we compared cobas®Liat® PCR System (Liat) and DIAs, and Liat showed a higher positive rate than DIAs (51.6% versus 40.7%), and the difference was evident within 18h from the onset of symptoms. [12] Since other previous studies also reported a higher sensitivity of molecular POC tests compared to DIAs,[2,13,14] molecular POC tests, including TRCsatFLU, can contribute to the accurate diagnosis of influenza.

There are several rapid RT-PCR technologies for the detection of influenza, such as Liat, ID NOW (formerly Alere™ i), and GeneXpert Xpress (Xpert). Some multicenter studies have reported their performance for detecting influenza: the sensitivity/specificity of Liat, ID NOW, and Xpert for influenza A and B was reported as 0.996/0.975 and 0.993/0.997, 0.978– 0.993/0.966–0.981 and 0.929–0.976/0.983–1.000, and 0.953–1.000/0.948–1.000 and 0.938-1.000/0.995-1.000. [15–20] Compared to the results of these previous studies, TRCsatFLU showed comparable sensitivity and specificity in nasopharyngeal swabs. TRCsatFLU has a unique feature: it can detect influenza in gargle samples with simple sample preparation without purification. In this study and previous single-center studies, TRCsatFLU showed great sensitivity and specificity in gargle samples.[5] Some studies have reported the performance of molecular POC tests on gargle samples, and the sensitivity of Xpert and Liat for influenza detection in gargle samples is 0.917 (11/12 samples) and 1.000 (15/15 samples), respectively, compared to in-house RT-PCR.[21,22] In addition to gargle samples, several non-nasopharyngeal swab specimens, such as nasal aspirate-wash, nasopharyngeal aspirate, throat swabs, and saliva have been evaluated using molecular POC tests. The sensitivity of molecular POC tests in the nasal aspirate wash, nasopharyngeal aspirate, throat swabs, and saliva was 0.900-1.000, 0.980-1.000, 0.75-0.83, and 0.750-0.918, respectively.[19,23,24] Among these samples, only nasopharyngeal aspirate showed high sensitivity comparable to gargle samples in this study.[23] The results in previous studies [5,19,21–24] and this study indicate that the gargle sample is an excellent specimen for detecting influenza by molecular POC tests. However, the sample preparation method was not described in the previous studies that evaluated the gargle samples using Xpert and Liat. [21,22] In addition, molecular POC tests other than TRCsatFLU do not officially designate gargle samples as specimens. Therefore, further studies are needed to determine whether gargle samples can be used as specimens in molecular POC tests other than TRCsatFlu.

In this study, the positive rate for influenza in TRCsatFLU was higher in the gargle samples (46.5%) than in nasopharyngeal swabs (41.6%). However, in the 202 patients for whom both samples were taken, the positive rate was slightly lower in gargle samples (45.5%) than in nasopharyngeal swabs (46.5%). Among the 202 patients, 8 were positive for influenza only in nasopharyngeal swabs, and 6 were positive for influenza only in gargle samples, but there were no significant differences in patient background between them. Therefore, it is not clear which cases of gargle samples should be used in preference to nasopharyngeal swabs. When compared to patients who tested positive for influenza in both samples, the influenza-positive rate by DIAs and RT-PCR and the percentage of diagnosing influenza were significantly lower in patients who tested positive for influenza only in either of the samples. These results suggest that the viral load in the sample was low in the patients who tested positive for influenza only in either of the samples.

This study had some limitations. We could only collect samples for one season because of the COVID-19 pandemic, as described in the Materials and Methods section. Therefore, we analyzed the data collected from fewer patients than initially planned. In addition, only one patient with an influenza B infection was included in this study. In a previous study, the TRCsatFLU showed high sensitivity and specificity for influenza B in nasopharyngeal swabs [5] but its performance in gargle samples remains unknown. In this study, TRCsatFLU has not been compared with other molecular POC tests. Thus, a comparative study is needed to determine if TRCsatFLU performs as well as other molecular POC tests.

## Conclusions

The novel molecular POC test, TRCsatFLU, showed great sensitivity and specificity for the detection of influenza. Since TRCsatFLU detects influenza in gargle samples collected by the patient, it can contribute to reducing the exposure risk to pathogens during sample collection by healthcare professionals.

## Supporting information

metadata file

## Data Availability

The derived data supporting the findings of this study are presented in this paper, supplementary Table1 and 2, and metadata file.

## Declarations

### Ethics approval

This study was approved by the ethics committee of Nagasaki University Hospital (approval number:19121603). : Before sample collection, written informed consent for the participation of this study was obtained from all participants.

### Consent for publication

Before sample collection, written informed consent for the publication of this study was obtained from all participants.

### Authors’ contributions

All the authors were involved in the study design. TU, TI, CY, YN, YO, KH, KH, and AT acquired the samples and data. NK, KI, HK, and KY were involved in data interpretation and analysis. NK wrote the original manuscript, and all authors revised and approved the manuscript for publication.

## Acknowledgements

none

## References

1. Chartrand C, Leeflang MMG, Minion J, Brewer T, Pai M. Accuracy of rapid influenza diagnostic tests: A meta-analysis. Ann Intern Med. American College of Physicians; 2012;156:500–11.

2. Merckx J, Wali R, Schiller I, Caya C, Gore GC, Chartrand C, et al. Diagnostic accuracy of novel and traditional rapid tests for influenza infection compared with reverse transcriptase polymerase chain reaction. Ann Intern Med. Ann Intern Med; 2017;167:395–409.

3. Drouillon V, Delogu G, Dettori G, Lagrange PH, Benecchi M, Houriez F, et al. Multicenter Evaluation of a Transcription-Reverse Transcription Concerted Assay for Rapid Detection of Mycobacterium tuberculosis Complex in Clinical Specimens. J Clin Microbiol. American Society for Microbiology Journals; 2009;47:3461–5.

4. Mazzarelli A, Cannas A, Venditti C, D’Arezzo S, De Giuli C, Truffa S, et al. Clinical evaluation of TRCReady M.TB for rapid automated detection of M. tuberculosis complex in respiratory samples. Int J Tuberc Lung Dis. Int J Tuberc Lung Dis; 2019;23:260–4.

5. Kaku N, Hashiguchi K, Akamatsu N, Wakigawa F, Matsuda J, Komaru K, et al. Evaluation of a novel rapid TRC assay for the detection of influenza using nasopharyngeal swabs and gargle samples. Eur J Clin Microbiol Infect Dis. Eur J Clin Microbiol Infect Dis; 2021;40:1743–8.

6. Kaku N, Ota K, Sasaki D, Akamatsu N, Uno N, Sakamoto K, et al. Had COVID-19 spread in the community before the first confirmed case in Nagasaki, Japan? Microbes Infect. Microbes Infect; 2021;23:104812.

7. Kaku N, Nishimura F, Shigeishi Y, Tachiki R, Sakai H, Sasaki D, et al. Performance of anti-SARS-CoV-2 antibody testing in asymptomatic or mild COVID-19 patients: A retrospective study in outbreak on a cruise ship. PLoS One. Public Library of Science; 2021;16:e0257452.

8. Ota K, Yanagihara K, Sasaki D, Kaku N, Uno N, Sakamoto K, et al. Detection of SARS-CoV-2 using qRT-PCR in saliva obtained from asymptomatic or mild COVID-19 patients, comparative analysis with matched nasopharyngeal samples. PLoS One. Public Library of Science; 2021;16:e0252964.

9. World Health Organization. World Health Organization Global Epidemiological Surveillance Standards for Influenza [Internet]. 2013. Available from: https://www.who.int/publications/i/item/9789241506601

10. Mitamura K, Shimizu H, Yamazaki M, Ichikawa M, Nagai K, Katada J, et al. Clinical evaluation of highly sensitive silver amplification immunochromatography systems for rapid diagnosis of influenza. J Virol Methods. 2013;194:123–8.

11. National Institute of Infectious Diseases. Manual for diagnosis of influenza [Internet]. 4th ed. Tokyo; 2019. p. 24–57. Available from: https://www.niid.go.jp/niid/images/lab-manual/influenza20190116.pdf

12. Kaku N, Kodama H, Akamatsu N, Ota K, Kosai K, Morinaga Y, et al. Multicenter evaluation of molecular point-of-care testing and digital immunoassays for influenza virus A/B and respiratory syncytial virus in patients with influenza-like illness. J Infect Chemother. Elsevier; 2021;27:820–5.

13. Kanwar N, Michael J, Doran K, Montgomery E, Selvarangan R. Comparison of the ID Now Influenza A & B 2, Cobas Influenza A/B, and Xpert Xpress Flu Point-of-Care Nucleic Acid Amplification Tests for Influenza A/B Virus Detection in Children. J Clin Microbiol. J Clin Microbiol; 2020;58.

14. Sato Y, Nirasawa S, Saeki M, Yakuwa Y, Ono M, Kobayashi R, et al. Comparative study of rapid antigen testing and two nucleic acid amplification tests for influenza virus detection. J Infect Chemother; 2022;28:1033–6.

15. Gibson J, Schechter-Perkins EM, Mitchell P, Mace S, Tian Y, Williams K, et al. Multi-center evaluation of the cobas® Liat® Influenza A/B & RSV assay for rapid point of care diagnosis. J Clin Virol. J Clin Virol; 2017;95:5–9.

16. Hassan F, Crawford J, Bonner AB, Ledeboer NA, Selvarangan R. Multicenter evaluation of the Alere™ i influenza A&B assay using respiratory specimens collected in viral transport media. Diagn Microbiol Infect Dis. Diagn Microbiol Infect Dis; 2018;92:294–8.

17. Bell J, Bonner A, Cohen DM, Birkhahn R, Yogev R, Triner W, et al. Multicenter clinical evaluation of the novel Alere™ i Influenza A&B isothermal nucleic acid amplification test. J Clin Virol. J Clin Virol; 2014;61:81–6.

18. Wolters F, Grünberg M, Huber M, Kessler HH, Prüller F, Saleh L, et al. European multicenter evaluation of Xpert® Xpress SARS-CoV-2/Flu/RSV test. J Med Virol. J Med Virol; 2021;93:5798–804.

19. Novak-Weekley S, Marlowe EM, Poulter M, Dwyer D, Speers D, Rawlinson W, et al. Evaluation of the Cepheid Xpert Flu Assay for rapid identification and differentiation of influenza A, influenza A 2009 H1N1, and influenza B viruses. J Clin Microbiol. J Clin Microbiol; 2012;50:1704–10.

20. Cohen DM, Kline J, May LS, Harnett GE, Gibson J, Liang SY, et al. Accurate pcr detection of influenza a/b and respiratory syncytial viruses by use of cepheid xpert flu+rsv xpress assay in point-of-care settings: Comparison to prodesse proflu+. J Clin Microbiol. J Clin Microbiol; 2018;56.

21. Bennett S, MacLean A, Gunson R. Verification of Cepheid Xpert Xpress Flu/RSV assay for use with gargle samples, sputa and endotracheal secretions. 2019;101:114–5.

22. Goldstein EJ, Gunson RN. In-house validation of the cobas Liat influenza A/B and RSV assay for use with gargles, sputa and endotracheal secretions. J Hosp Infect. J Hosp Infect; 2019;101:289–91.

23. To KKW, Yip CCY, Lai CYW, Wong CKH, Ho DTY, Pang PKP, et al. Saliva as a diagnostic specimen for testing respiratory virus by a point-of-care molecular assay: a diagnostic validity study. Clin Microbiol Infect. Clin Microbiol Infect; 2019;25:372–8.

24. Davis S, Allen AJ, O’Leary R, Power M, Price DA, Simpson AJ, et al. Diagnostic accuracy and cost analysis of the Alere™ i Influenza A&B near-patient test using throat swabs. J Hosp Infect. J Hosp Infect; 2017;97:301–9.

25. Frazee BW, Rodríguez-Hoces de la Guardia A, Alter H, Chen CG, Fuentes EL, Holzer AK, et al. Accuracy and Discomfort of Different Types of Intranasal Specimen Collection Methods for Molecular Influenza Testing in Emergency Department Patients. Ann Emerg Med. Ann Emerg Med; 2018;71:509-517.e1.

26. Malecki M, Lüsebrink J, Teves S, Wendel AF. Pharynx gargle samples are suitable for SARS-CoV-2 diagnostic use and save personal protective equipment and swabs. Infect. Control Hosp. Epidemiol. Infect Control Hosp Epidemiol; 2021. p. 248–9.

